# Equitable Artificial Intelligence for Glaucoma Screening with Fair Identity Normalization

**DOI:** 10.1101/2023.12.13.23299931

**Authors:** Min Shi, Yan Luo, Yu Tian, Lucy Shen, Tobias Elze, Nazlee Zebardast, Mohammad Eslami, Saber Kazeminasab, Michael V. Boland, David S. Friedman, Louis R. Pasquale, Mengyu Wang

## Abstract

**Objective:** To develop an equitable artificial intelligence model for glaucoma screening.

**Design:** Cross-sectional study.

**Participants:** 7,418 optical coherence tomography (OCT) paired with reliable visual field (VF) measurements of 7,418 patients from the Massachusetts Eye and Ear Glaucoma Service between 2021 and 2023.

**Methods:** We developed fair identify normalization (FIN) module to equalize the feature importance across different identity groups to improve model performance equity. EfficientNet served as the backbone model to demonstrate the effect of FIN on model equity. The OCT-derived retinal nerve fiber layer thickness (RNFLT) maps and corresponding three-dimensional (3D) OCT B-scans were used as model inputs, and a reliable VF tested within 30 days of an OCT scan was used to categorize patients into glaucoma (VF mean deviation < -3 dB, abnormal glaucoma hemifield test (GHT) and pattern standard deviation (PSD) < 5%) or non-glaucoma (VF mean deviation ≥ -1 dB and normal GHT and PSD results). The area under the receiver operating characteristic curve (AUC) was used to measure the model performance. To account for the tradeoff between overall AUC and group disparity, we proposed a new metric called equity-scaled AUC (ES-AUC) to compare model performance equity. We used 70% and 30% of the data for training and testing, respectively.

**Main Outcome Measures:** The glaucoma screening AUC in different identity groups and corresponding ES-AUC.

**Results:** Using RNFLT maps with FIN for racial groups, the overall AUC and ES-AUC increased from 0.82 to 0.85 and 0.76 to 0.81, respectively, with the AUC for Blacks increasing from 0.77 to 0.81. With FIN for ethnic groups, the overall AUC and ES-AUC increased from 0.82 to 0.84 and 0.77 to 0.80, respectively, with the AUC for Hispanics increasing from 0.75 to 0.79. With FIN for gender groups, the overall AUC and ES-AUC increased from 0.82 to 0.84 and 0.80 to 0.82, respectively, with an AUC improvement of 0.02 for both females and males. Similar improvements in equity were seen using 3D OCT B scans. All differences regarding overall-and ES-AUCs were statistically significant (p < 0.05).

**Conclusions:** Our deep learning enhances screening accuracy for underrepresented groups and promotes identity equity.

## Introduction

Glaucomatous optic neuropathy is the leading cause of irreversible blindness globally^1–4^ affecting 3.5% of the population between 40 and 80 years totaling 3 million patients in the US and 80 million patients worldwide.^1,5^ However, most commonly, glaucoma patients are not aware of having the disease until the vision loss becomes severe enough to impair their daily activities, such as reading and driving due to the brain and fellow eye compensation.^6–12^ It has been reported that 50% of people with glaucoma do not know they have the disease, and racial and ethnic minority groups are particularly affected due to a lack of access to ophthalmic care largely attributed to financial limitations.^13,14^

Increasing research indicates that glaucoma disproportionately impacts racial and ethnic minorities and socioeconomically disadvantaged identity groups.^15–21^ We recently found that visual field (VF) loss in glaucoma patients at the first visit to an ophthalmology service is significantly worse in Blacks and Asians than in Whites and significantly worse in Hispanics than in non-Hispanics.^22^ Noticeably, lower proficiency in English was also linked to a more pronounced VF loss in patients with glaucoma. Despite greater severity at the first visit, Black patients had lower VF test frequency compared to Whites. In a separate study, we found that compared with non-Hispanic Whites, Black individuals faced higher risks of developing early central and advanced VF loss.^23^ Blacks and Hispanics are significantly more likely (4.4 and 2.5 times) to have undiagnosed and untreated glaucoma compared with non-Hispanic Whites.^13^ Therefore, automated glaucoma screening with deep learning deployed at primary care and pharmacies would greatly benefit racial and ethnic minorities and socioeconomically disadvantaged identity groups.

Though numerous deep learning studies have been conducted for automated glaucoma detection using retinal images (e.g. as illustrated in **Figure 1a**),^24–31^ it remains unclear if these deep learning models have equitable performance across different identity groups. In recent years, significant work in deep learning has been done to alleviate performance inequality in the models.^32–34^ The performance inequality observed in deep learning models primarily stems from data inequality and data characteristic variability between different identity groups. Standard deep learning models without equity-improving design favor the majority group algorithmically and may not be able to represent the diverse data characteristic variability across different identity groups. For example, consistent with the US population composition,^35^ there are fewer Black and Asian glaucoma patients present in ophthalmic care, which is data inequality.^36^

**Figure 1:**
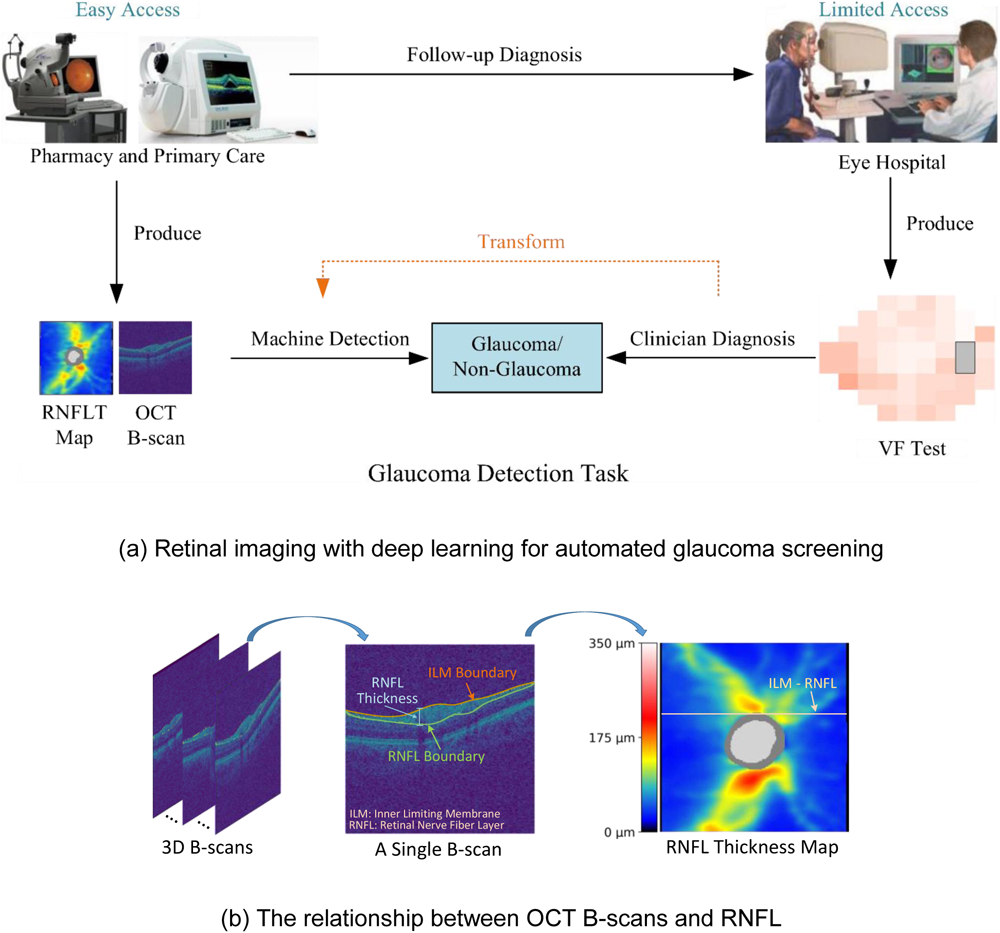
The glaucoma screening paradigm with RNFLT maps and 3D B-scans. **(a)** Illustration of using retinal imaging with deep learning for automated glaucoma screening, and **(b)** the relationship between OCT B-scans and retinal nerve fiber layer (RNFL). OCT: optical coherence tomography; VF: visual field; RNFLT: RNFL thickness.

In this study, we introduce fair identity normalization (FIN) to promote equitable glaucoma screening. The fundamental premise of FIN rests on the notion that individuals within the same identity group exhibit a greater correlation compared to those from other groups. This correlation is cultivated by promoting distinct feature distributions across different identity groups during the deep learning model’s training phase. Our examination of FIN’s efficacy spanned across two state-of-the-art deep learning frameworks: EfficientNet and ResNet.^37,38^ However, given that EfficientNet is more effective than ResNet based on our experiments, we chose EfficientNet as the backbone model in this study. We aimed to diminish group disparities in glaucoma screening using retinal nerve fiber layer thickness (RNFLT) maps. Additionally, a three-dimensional (3D) convolutional neural network (CNN),^39^ both standalone and in conjunction with FIN, was employed to predict glaucomatous status using 3D optical coherence tomography (OCT) B-scans. We compared FIN against other common methods. These included oversampling to equalize data representation from various groups, and a transfer learning approach where a deep learning model trained on the entire patient data was fine-turned for each individual identity group based on race, gender, and ethnicity, respectively. We adopted the area under the receiver operating characteristic curve (AUC) to analyze overall screening accuracy and group-level accuracies. Furthermore, to account for the tradeoff between overall AUC and group disparity, we proposed a new metric called equity-scaled AUC (ES-AUC) to compare the model equity. Additionally, we used mean and max disparities to quantify the differences in screening accuracies across different identity groups.

## Methods

The OCT data used for developing the equitable deep learning model were from the glaucoma patient service at the Massachusetts Eye and Ear (MEE) between 2021 and 2023. The institutional review boards (IRB) of MEE approved the creation of the database in this retrospective study. This study complied with the guidelines outlined in the Declaration of Helsinki. In light of the study’s retrospective design, the requirement for informed consent was waived.

### Dataset Description

In this study, we utilized a dataset comprising 7,418 RNFLT maps obtained from 7,418 patients who underwent tests at the MEE glaucoma service from 2021 to 2023. Each of these two-dimensional (2D) RNFLT maps (**Figure 1b**), with dimensions of 200 × 200, represents thickness values and was sourced from a spectral-domain OCT instrument (Cirrus, Carl Zeiss Meditec, Dublin, California). Only high-quality RNFLT maps with a signal strength of 6 or higher were considered. Additionally, the dataset encompassed corresponding 3D OCT B-scans. Each of these 3D OCT B-scan volumes consists of 200 individual B-scans, with each B-scan measuring 200 × 200 in dimension.

The glaucomatous status was determined by matching the reliable visual field (VF) test with OCT. In this study, we exclusively utilized reliable 24-2 VFs, characterized by fixation loss ≤ 33%, a false positive rate ≤ 20%, and a false negative rate ≤ 20%. These reliability criteria align with those employed in our previous research.^40,41^ Glaucomatous status was ascertained based on the VF mean deviation (MD): an MD of less than -3 dB with abnormal glaucoma hemifield test and pattern standard deviation results was classified as glaucoma, while an MD greater than or equal to -1 dB with normal glaucoma hemifield test and pattern standard deviation results was identified as non-glaucoma.

### Glaucoma Screening Model with Fair Identity Normalization

We designed a deep learning model based on the EfficientNet,^37^ enhanced with fair identity normalization (FIN), to achieve equitable glaucoma screening (**Figure 2**). Initially, the model takes RNFLT maps or 3D OCT B-scans as the input, extracting pertinent and discriminative features through the EfficientNet structure. Subsequently, these features undergo normalization via FIN, considering the group identity associated with the input image. As a result of this normalization, data from patients within the same identity groups are aligned to a consistent distribution, thereby aiding the model in distinguishing between different identity groups. Finally, the normalized features, tailored to specific identity groups, are employed to predict glaucomatous status. The model training followed a supervised approach. The dataset underwent a patient-level random split: 70% was dedicated to model training and the remaining 30% was used for evaluating both glaucoma screening precision and group equity.

**Figure 2:**
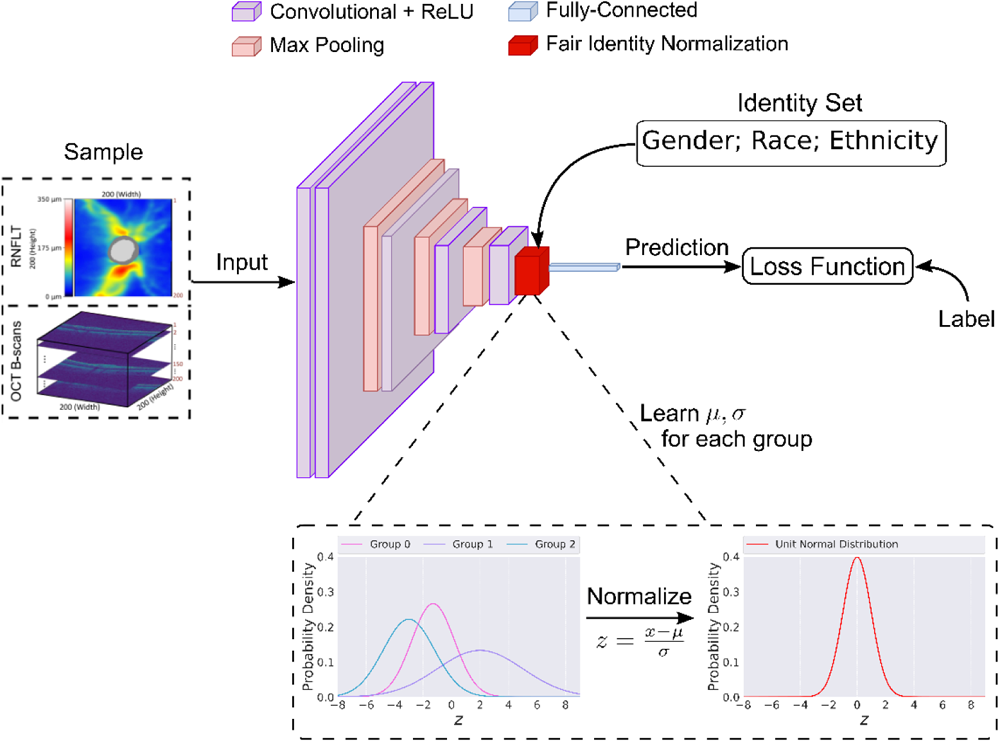
The proposed far identity normalization approach for equitable glaucoma screening.

We assessed FIN’s efficacy for equitable glaucoma screening across three distinct identity parameters: race, ethnicity, and gender. Beyond EfficientNet, we also examined the effectiveness of FIN through its integration with ResNet for glaucoma detection. Furthermore, we benchmarked EfficientNet + FIN against oversampling and transfer learning methods, known to be helpful to mitigate group disparities which could be caused by the skewed distribution of samples between different identity groups.

### Evaluation of Glaucoma Screening and Group Disparity

We employed deep learning models, both with and without FIN integration, to predict glaucomatous status. The AUC served as our metric for quantifying both overall screening accuracy and group-level accuracy, categorized by race, gender, and ethnicity. Specifically, we concentrated on the racial groups of Asians, Blacks, and Whites; the gender-based groups of Females and Males; and the ethnic groups of Non-Hispanics and Hispanics. To ascertain AUC disparities in glaucoma screening across these identity groups, we proposed to use a new metric called equity-scaled AUC (ES-AUC) to compare the model performance equity. ES-AUC was computed as the ratio of the overall AUC to the adjusted (incremented by one) sum of differences between the overall AUC and every individual group AUC. Additionally, we calculated both mean disparity and max disparity. Mean disparity was determined by the ratio of the standard deviation of individual group AUCs to the adjusted overall AUC, which is the overall AUC subtracting 0.5. Meanwhile, max disparity was computed as the ratio of the difference between the highest and lowest individual group AUCs to the same adjusted overall AUC.

We used paired t-test and bootstrapping with replacement to compare the glaucoma screening performance of different deep learning models with or without FIN. All deep learning modeling and statistical analyses were performed in Python 3.8 (available at http://www.python.org) on a Linux system.

## Results

### Participant Characteristics

In this study, we analyzed 3D OCT B-scans and their corresponding RNFLT maps from 7,418 eyes of 7,418 unique patients. The participants’ average age at the time of imaging was 60.8 ± 16.5 years, and 57.8% were females. When examining ethnicity and race, 4.6% of the patients identified as Hispanic, while the racial distribution was 8.6% Asian, 14.9% Black, and 76.5% White (as shown in **Table 1**). Notably, 46.7% of these 7,418 patients were diagnosed with glaucoma.

**Table 1:** Demographics and baseline characteristics of the patients included in the evaluation.

| Characteristic | N= patients |
| --- | --- |
| Number of patients (eyes) | 7,418 (7,418) |
| Number of 3D OCT B-scans and RNFLT map pairs | 7,418 |
| Age (years) | 60.8 ± 16.5 |
| Gender (female %) | 57.8 |
| Prevalence of right eyes (%) | 47.2 |
| Prevalence of Hispanic (%) | 4.6 |
| Race (%) |  |
| • Asian | 8.6 |
| • Black or African American | 14.9 |
| • White or Caucasian | 76.5 |
| Visual field parameter |  |
| • Average HVF MD (dB) | -4.0 ± 5.9 |
| • Average TD (dB) | -4.1 ± 6.0 |
| Prevalence of glaucoma (%) | 46.7% |

Age is presented as mean ± standard deviation, unless otherwise stated.

### Glaucoma Screening Performance using RNFLT Maps

For ResNet combined with FIN, the overall AUC and ES-AUC for racial group increased from 0.76 and 0.80 to 0.77 and 0.82 (p-value < 0.001), with the AUC improved by 0.01 (p-value < 0.05) for Blacks, and 0.02 (p-value < 0.001) for both Asians and Whites, although the improvements (from 0.09 and 0.17 to 0.08 and 0.17) of mean and max disparities are not significant (**Figure 3a**). Similarly, the overall AUC and ES-AUC for gender group both improved by 0.02, and the mean and max disparities increased from 0.09 and 0.12 to 0.06 and 0.09 (p-value < 0.001), respectively (**Figure 3b**). For ethnic group, the overall AUC had an improvement of 0.02 (p-value < 0.001), while the ES-AUC remained unchanged after integrating FIN with ResNet (**Figure 3c**).

**Figure 3:**
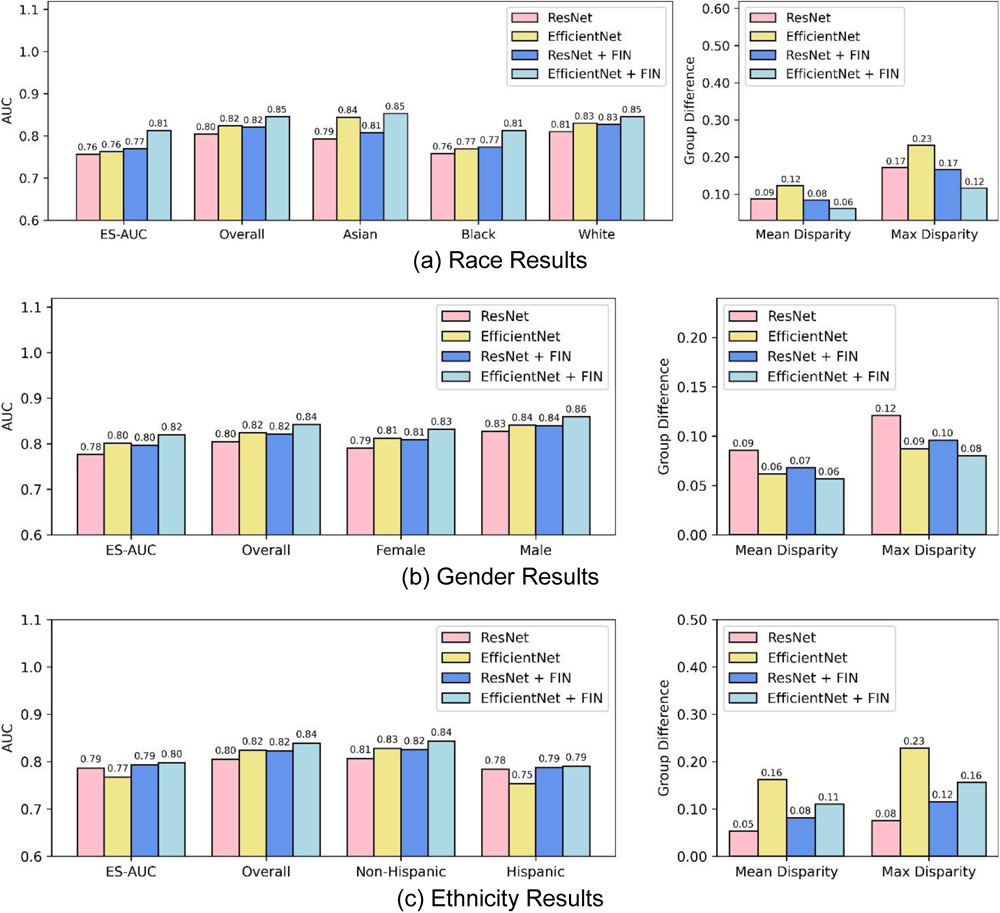
Comparison of EfficientNet and ResNet using retinal nerve fiber layer thickness maps from optical coherence tomography scans for glaucoma detection across different demographic identity groups. **(a)** race results, **(b)** gender results, and **(c)** ethnicity results. FIN: fair identity normalization.

In comparison, after combining the FIN with EfficientNet, the overall AUC and ES-AUC for racial groups increased from 0.82 to 0.85 and 0.76 to 0.81 (p-value < 0.001), respectively, with the AUC for Blacks increasing from 0.77 to 0.81 (p-value < 0.001) (**Figure 3a**). The mean and max disparities significantly decreased from 0.12 to 0.06 and 0.23 to 0.12 (p-value < 0.001) (**Figure 3a**), respectively. With FIN for gender groups, the overall AUC and ES-AUC increased from 0.82 to 0.84 and 0.80 to 0.82 (p-value < 0.001), respectively, with an AUC improvement of 0.02 for both females and males (p-value < 0.001) (**Figure 3b**). With FIN for ethnic groups, the overall AUC and ES-AUC increased from 0.82 to 0.84 and 0.77 to 0.80 (p-value < 0.001), respectively, with the AUC for Hispanics increasing from 0.75 to 0.79 (p-value < 0.001) (**Figure 3c**). The mean and max disparities decreased by 0.05 and 0.07 (p-value < 0.001), respectively (**Figure 3c**).

The feature distributions learned by the deep learning model from input RNFLT maps are shown in **Figure 4**. With FIN for racial groups, features of Asians and Blacks were more deviant, and they both are closer to the features of Whites (**Figure 4a** and **Figure 4d**). For gender groups, the features of females and males are more similar after the integration of FIN (**Figure 4b** and **Figure 4e**). In contrast, the features between non-Hispanic and Hispanic groups became more similar with FIN (**Figure 4c** and **Figure 4f**).

**Figure 4:**
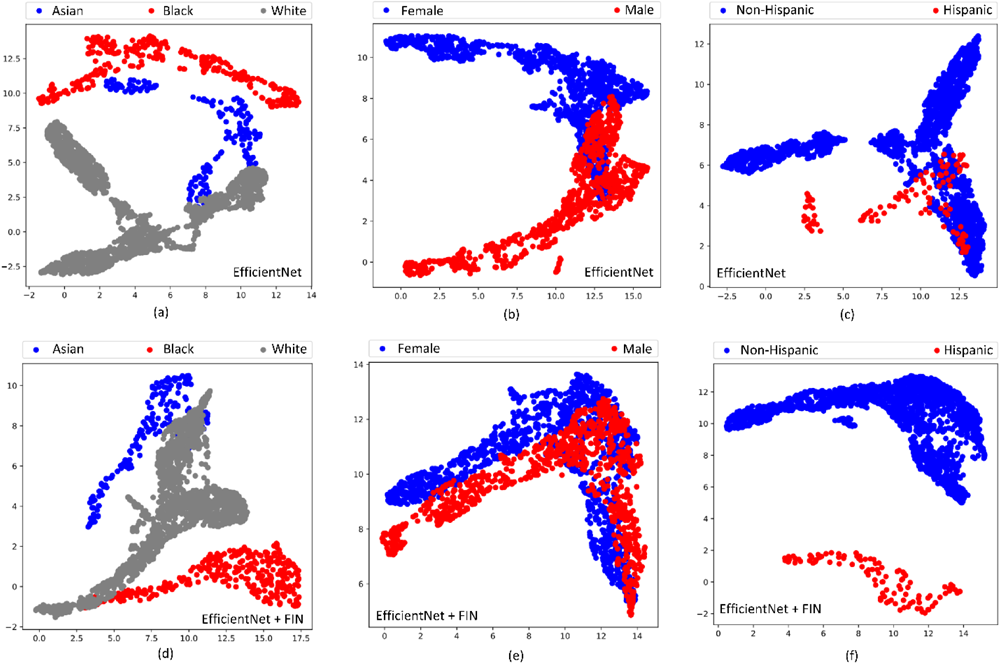
UMAP-generated distribution of features learned from RNFLT maps. (a) Feature distribution in racial groups with EfficientNet. (b) Feature distribution in gender-based groups with EfficientNet. (c) Feature distribution in ethnic groups with EfficientNet. (d) Feature distribution in racial groups with EfficientNet + FIN. (e) Feature distribution in gender-based groups with EfficientNet + FIN. (f) Feature distribution in ethnic groups with EfficientNet + FIN.

While comparing the oversampling and transfer learning methods, FIN generally achieved better AUC and ES-AUC performances for different identity groups (**Figure 5**). For racial groups, the overall AUC and ES-AUC of FIN were 0.03 and 0.04 higher than the oversampling (p-value < 0.001), and 0.01 and 0.04 higher than the transfer learning (p-value < 0.001) (**Figure 5a**). The mean and max disparities of FIN had a significant decrease of 0.04 and 0.10 compared to the over sampling approach (p-value < 0.001), and 0.07 and 0.15 compared to the transfer learning (p-value < 0.001). Similarly, for gender groups, the overall AUC and ES-AUC of FIN both improved by 0.05 compared with oversampling (p-value < 0.001), where the improvements were 0.01 and 0.04 compared to the transfer learning (p-value < 0.001) (**Figure 5b**). The mean and max disparities of FIN significantly decreased by 0.02 and 0.03 compared to oversampling (p-value < 0.001), and 0.07 and 0.1 compared to the transfer learning (p-value < 0.001). Lastly for ethnic groups, the overall AUC and ES-AUC of FIN significantly improved by about 0.02 and 0.01 over the sampling and transfer learning methods (p-value < 0.001), respectively (**Figure 5c**).

**Figure 5:**
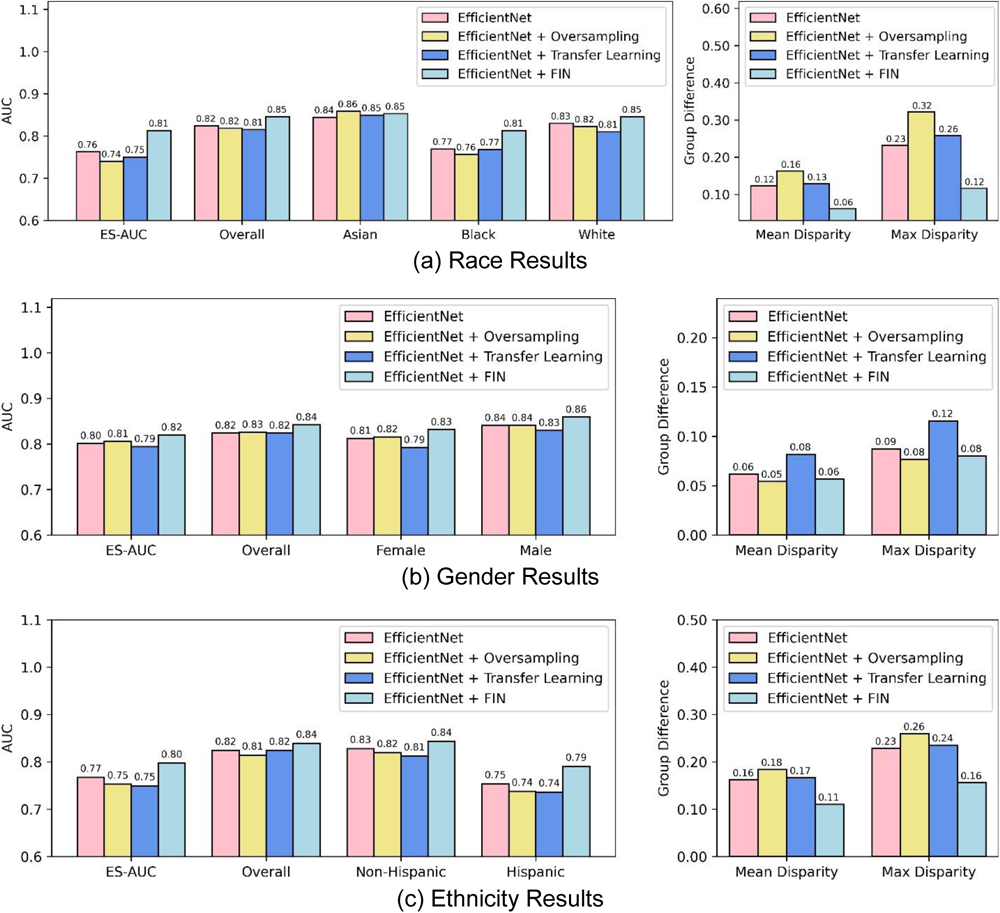
Comparison of various methods using retinal nerve fiber layer thickness maps from optical coherence tomography scans for glaucoma detection across different demographic identity groups. **(a)** race results, **(b)** gender results, and **(c)** ethnicity results. FIN: fair identity normalization.

### Glaucoma Screening Performance using 3D OCT B-Scans

Using 3D OCT B-scans with FIN for glaucoma screening, the overall AUC and ES-AUC for racial groups improved from 0.84 to 0.85 and 0.78 to 0.80 (p-value < 0.05), respectively (**Figure 6a**). The mean and max disparities decreased by 0.01 and 0.03, respectively. With FIN for gender groups, the overall AUC and ES-AUC both had a moderate improvement of 0.01 (**Figure 6b**). While with FIN for ethnic group, the overall AUC and ES-AUC improved by 0.01 and 0.03 (p-value < 0.05), respectively, with the AUCs improved by 0.02 and 0.04 for non-Hispanic and Hispanic groups (p-value < 0.001) (**Figure 6c**). In addition, the mean and max disparities had significant declines of 0.06 and 0.08 (p-value < 0.001), respectively.

**Figure 6:**
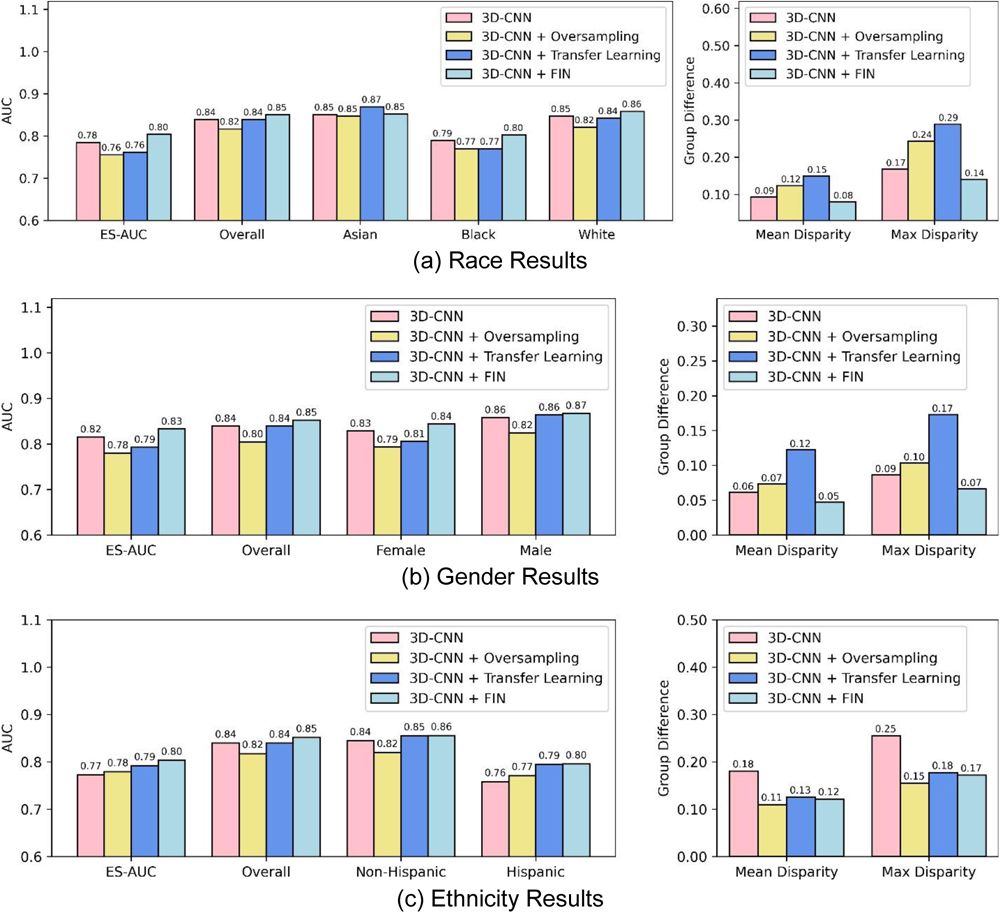
Comparison of various methods using three-dimensional optical coherence tomography scans for glaucoma detection across different demographic identity groups. **(a)** race results, **(b)** gender results, and **(c)** ethnicity results. FIN: fair identity normalization.

## Discussion

This paper is to demonstrate that deep-learning models for glaucoma screening may perform quite differently across demographic groups, and it is possible to reduce the performance disparity gap between different demographic groups by model innovation, which is our fair identity normalization (FIN) model in this work. Simply using our fair identity normalization model without any additional real cost can make deep-learning glaucoma screening models more equitable, which means reduced group disparities with no overall performance deterioration or even overall performance improvement.

In this study, we introduced a deep learning model that combines EfficientNet with FIN for equitable glaucoma screening utilizing RNFLT maps and 3D OCT B-scans (**Figure 2**). In comparison to the standalone EfficientNet, the EfficientNet combined with FIN model enhanced the overall AUC by 0.03 and ES-AUC by 0.05 for racial groups, with noticeable improvements in AUCs when delineated by race, ethnicity, and gender. We have compared EfficientNet with another state-of-the-art deep learning model ResNet with and without the enhancement of FIN (**Figure 3**). Both deep learning models showed improved glaucoma screening performance and equity with FIN, which demonstrates that FIN is effective in promoting equitable glaucoma screening.

Previous studies have consistently highlighted that glaucoma has a disproportionate impact on racial and ethnic minorities, as well as socioeconomically marginalized groups. For instance, upon their initial visit to an ophthalmology service, Black and Asian glaucoma patients typically display more severe visual field (VF) loss than their White counterparts. Similarly, the severity is worse in Hispanic patients compared to non-Hispanics. Notably, Blacks and Hispanics are approximately 4.4 and 2.5 times more likely, respectively, to have undetected and untreated glaucoma than non-Hispanic Whites. While deep learning models have gained traction for automated glaucoma detection, they often overlook the crucial aspect of ensuring equal performance across diverse identity groups. Our proposed FIN effectively addresses these disparities across different identity groups. For example, using RNFLT maps for glaucoma screening with FIN, racial groups increased from 0.82 to 0.85 and 0.76 to 0.81, respectively, with the AUC for Blacks increasing from 0.77 to 0.81 (**Figure 3a**). In addition, there was a significant decrease in the mean and max disparities by 0.06 and 0.11 for Asian, Black, and White groups (**Figure 3a**). The incorporation of FIN with ResNet, as well as its comparison to oversampling and transfer learning techniques, further underscores the efficacy of FIN in fostering equitable glaucoma screening.

Feature distribution visualizations indicate that FIN increases the distinction of learned features between Asians and Blacks compared to Whites (**Figure 4a** and **Figure 4d**). This suggests that the overlap of features for Asians and Blacks might have led to higher false positive rates in glaucoma screening for these groups. FIN’s enhancement of feature differentiation has sharpened the distinction, improving the AUC for Blacks by 0.04 (**Figure 3a**). Additionally, FIN has narrowed the feature gap between genders (**Figure 4b** and **Figure 4e**) and widened the feature variance between non-Hispanic and Hispanic groups. These changes demonstrate FIN’s role in refining feature distributions to optimize glaucoma screening outcomes and fairness.

Our research has several limitations. Firstly, even though we have achieved equitable glaucoma screening with comprehensive evaluations encompassing all severity stages, we have not examined the performance across different stages like mild versus severe glaucoma. This omission is significant since different identity groups could experience varied screening accuracies based on the stage of their condition. Secondly, while the patient distribution across identity groups in our study mirrors real-world disparities (for instance, 8.6%, 14.9%, and 76.5% were identified as Asian, Black, and White respectively), we have not assessed the equity of glaucoma screening in scenarios where sample sizes across these groups are balanced. Thirdly, our evaluation of FIN concentrated on its integration with EfficientNet and ResNet. We have not explored its efficacy when paired with other prevalent deep learning models like the vision transformer or the VGG network, even though FIN has the versatility to be paired with various learning frameworks. Lastly, our use of ES-AUC, mean and max disparities to measure the fairness of glaucoma prediction among identity groups is limited. There exists a range of other fairness metrics, such as demographic parity, equalized odds, and equal opportunity, which we have not considered.

While FIN typically aids in diminishing the mean and max disparities for race and gender, its integration with ResNet on RNFLT maps did not show any remarkable improvement in equity between the non-Hispanic and Hispanic groups ethnic. This observation can be linked to a couple of key factors. Firstly, the efficiency of FIN can vary based on the deep learning model it is paired with. Different models have unique capabilities in extracting beneficial features from the RNFLT map that can enhance equity. Secondly, the metrics we employed to measure the equity of glaucoma screening across various identity groups might not capture the full picture. To get a more holistic understanding, it would be beneficial to consider other fairness metrics in future research, such as demographic parity, equalized odds, and equal opportunity.

In summary, we introduced FIN that can seamlessly integrate with many mainstream deep learning frameworks for equitable glaucoma screening. FIN works by normalizing features derived from RNFLT maps or 3D OCT B-scans in alignment with patients’ identity groups. Our evaluations on prominent deep learning architectures, EfficientNet and ResNet, underscore FIN’s capability to not only boost glaucoma screening accuracy but also curtail disparities across race, ethnicity, and gender, particularly in conjunction with EfficientNet. When compared against other strategies like oversampling and transfer learning that aim for equity in glaucoma screening, FIN consistently outperforms. With its potential for real-world clinical applications, our deep learning model incorporating FIN stands as a promising tool to ensure equitable glaucoma screening outcomes across diverse identity groups.

## Data Availability

In accordance with the policies of Massachusetts Eye and Ear, the data from this study cannot be publicly disclosed

